# Health losses attributed to anthropogenic climate change

**DOI:** 10.1101/2024.08.07.24311640

**Authors:** Colin J. Carlson, Dann Mitchell, Rory Gibb, Rupert F. Stuart-Smith, Tamma Carleton, Torre E. Lavelle, Catherine A. Lippi, Megan Lukas-Sithole, Michelle A. North, Sadie J. Ryan, Dorcas Stella Shumba, Matthew Chersich, Mark New, Christopher H. Trisos

## Abstract

Over the last decade, health impact attribution studies have shown that climate change is a present-day public health emergency, with substantial impacts felt through death, disability, and illness, equivalent to financial losses on the order of US$ trillions. However, these studies have so far been biased towards the direct effects of heat and extreme weather in high-income countries, and so capture a small fraction of the total global burden of climate change. Expanding the use of attribution science in public health could help put pressure on policymakers to take action for human health.

---

It is unequivocal that recent climate change is outside the realm of normal natural variability, that natural factors in the earth system cannot explain the observed changes, and that anthropogenic influences are responsible for – at the time of writing – roughly +1.3°C of global warming from pre-industrial levels^1^. Climate scientists have developed this consensus through a set of quantitative methods that are grouped under the joint umbrella of *climate change detection* (showing that the climate has changed) and *attribution* (distinguishing the relative contributions of both anthropogenic and natural influences on the global climate system). In the last decade, researchers have also started using the same methods to isolate the effects of anthropogenic forcings on the social and ecological consequences of climate change. These *end-to-end impact attribution* studies^2^ account for a small fraction of total research on climate change impacts, but represent some of the strongest evidence in terms of both methodological rigor and ability to articulate clear, quantitative estimates of historical and present-day impacts.

Human health – especially loss of life, but also illness, disability, and poor well-being – is one of the most visible categories of climate change impacts. However, most work on the health impacts of climate change has stopped a step short of end-to-end attribution, focusing on long-term trends in health outcomes and their relationship to temperature and precipitation, or on the health outcomes of specific extreme weather events^3–5^. The first end-to-end *health impact attribution* study, which estimated the contribution of human-caused climate change to heat-related mortality during the 2003 European heat wave, was conducted in 2016^6^. Over the last decade, at least 20 studies have estimated the present-day health impacts of human-caused climate change (**Figure 1**; see **Online Methods** for search criteria).

**Figure 1.**
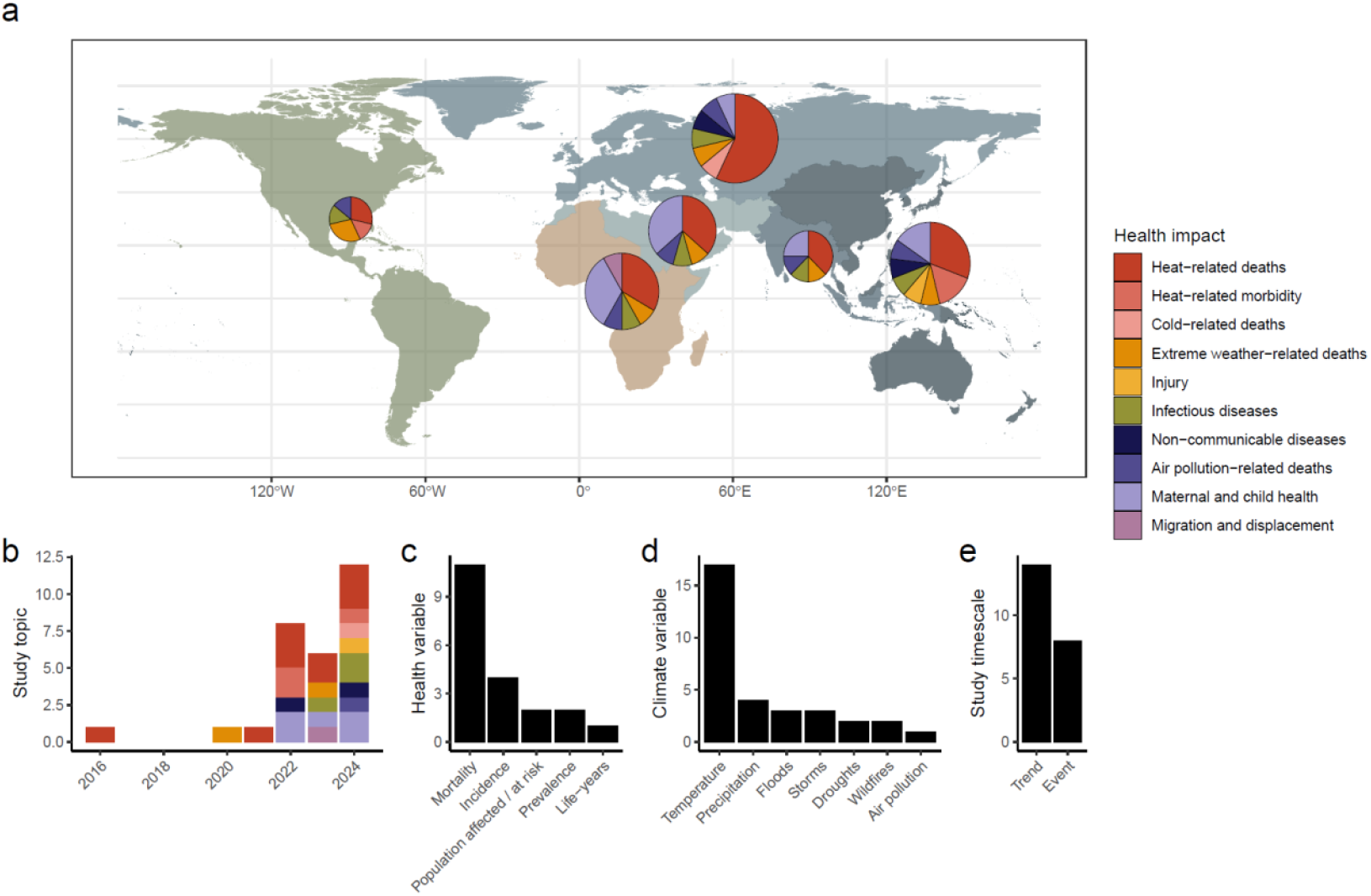
The rise of health impact attribution. (A) Distribution of research effort by health impact and geography of the affected population. The map is divided by World Health Organization global regions. Circle size is proportional to the number of applicable studies focused on each region, ranging from 7 in the Americas to 14 in Europe. (B-E) Distribution of studies across years (B) and study focus (C-E). Some studies are in multiple categories.

With a single exception^7^, every health impact attribution study to date has reported a substantial negative health impact of climate change – most often, a significant loss of life due to rising temperatures or extreme weather. Estimates of community-level mortality have ranged from 10 deaths (on a single day of the 2006 heat wave in London^8^) to 1,683 deaths (associated with heat in Zürich between 1969 and 2018^9^); at the national level and above, mortality estimates have ranged from 370 deaths (associated with the summer 2022 heat wave in Switzerland^10^) to 271,656 deaths (associated with heat across 43 countries between 1991 and 2018^11^) (**Table S1**). So far, research effort has been heavily biased towards temperature-related risks (n = 17 of 20), mortality (n = 11 of 20), and extreme weather events in Europe (n = 4 of 6). However, these studies have diversified over time, with recent research addressing the growing burden of mosquito-borne viral diseases^12,13^, mortality due to air pollution from wildfires^14^, population displacement by floods^15^, and several unique health risks experienced by children, including neonatal deaths^16,17^, preterm births (and life-long associations with asthma, type 1 and 2 diabetes, and cognitive disabilities)^17^, low birth weight^18^, and childhood malaria^19^.

In some cases, the estimated economic impact or value of these losses has been substantial. For example, one study estimated that medical costs related to heat wave-related preterm births in China could exceed US $300m per year, and the loss of lifetime earnings associated with cognitive disabilities could exceed US $1B per year^17^. Another study estimated that life-years lost due to Hurricane Harvey could be worth roughly US $17B^20^, while a global study of 185 extreme weather events estimated an average attributable loss of life valued at US $22.7B per year^21^. Applying the same standard estimates of the value of statistical life^22^, even with adjustment for national differences in GDP, suggests that annual losses due to some other attributable health impacts are also on the order of at least US $10B (see **Online Methods**). For example, we estimate attributable annual adjusted losses equivalent to US $17.1B (unadjusted: $106B) due to temperature-related neonatal deaths in 29 low- and middle-income countries^16^; $17.9B (unadjusted: $111.6B) due to heat-related deaths across 43 countries^11^; and US $23.2B (unadjusted: US $144.5B) due to global PM2.5 air pollution from wildfires[Citation error]. In some cases, directly quantified losses could be approaching the trillions: for example, Vicedo-Cabrera *et al*.’s estimate of 271,656 climate change-attributable heat-related deaths across 43 countries between 1991 and 2018 would be equivalent to a loss of US $502.6B (unadjusted: US $3.1T)^11^. These kinds of estimates are likely to be increasingly important as countries seek financing for loss and damage resulting from climate change^23^, particularly given that human health impacts dominate estimates of aggregate economic damages from future climate change^24^.

Another important frontier for health impact attribution is *source attribution*, a set of methods that quantify the contributions of specific major greenhouse gas emitters to extreme events and long-term warming trends, and in some cases, downstream impacts. For example, a recent preprint^9^ estimated that dozens of heat-related deaths in Switzerland between 1969 and 2018 could be attributed to specific fossil fuel companies, led by Chevron (59 deaths), ExxonMobil (54 deaths), and Saudi Aramco (53 deaths). Using the same value of statistical life approach as above, these estimates would be equivalent to losses of US $109.2m (unadjusted: $678.5m), US $99.9m (unadjusted: US $621m), and US $98.1m (unadjusted: US $609.5m) attributable to each emitter, respectively. Source attribution studies are likely to have a unique relevance to legal actions against emitters and governments, which can be supported by evidence linking anthropogenic greenhouse gas emissions – and potentially, the specific emissions of the defendant company – to the claimants’ losses^25,26^ (**Box 1**).

Increasingly, these studies represent the strongest available line of evidence regarding the present-day health impacts of climate change. In many cases, they are already substantially more up-to-date than modeling-based estimates: the only comprehensive estimate of present-day (circa 2000) global mortality and morbidity due to climate change was published twenty years ago^27,28^; in 2014, these estimates were extended from 2030 through 2050^29^. For some health impacts, such as heat- and extreme weather-related mortality, attribution studies have been broadly concordant with these projections (**Table 2**). In other cases, the divergence has been notable: for example, climate change-attributable mortality related to dengue fever has been an order of magnitude greater than expected, while malaria mortality is estimated to be an order of magnitude lower (see **Online Methods**). Some major expected sources of mortality have still not been re-assessed. For example, one attribution study estimated that every 1°C of anthropogenic warming has resulted in a 1-2% increase in food insecurity^30^, but no estimate exists of attributable mortality from malnutrition. Similarly, there have been no impact attribution studies focused on diarrheal diseases, despite several closely-related observational studies^31–33^.

New health impact attribution research continues to be published^34,35^. As the field of health impact attribution continues to grow, these studies can provide a more comprehensive view of the impacts of climate change on mortality, morbidity, life expectancy, and well-being^5,36^. In particular, future studies could assess the impact of climate change on dozens of infectious diseases; non-communicable diseases, including asthma, cancer, diabetes, heart disease, and kidney disease^37^; health impacts of food insecurity, including malnutrition, stunting, and direct mortality^30^; and mental health, including anxiety, depression, and suicides^38^. Future studies should also explore the uneven impacts of climate change within populations. These analyses are currently rare in the attribution literature, although one study found that women and the elderly accounted for 60% and 90% of attributable heat-related deaths, respectively, with older women experiencing mortality at 1.8 times the rate of the general population^10^.

Finally, future studies should aim to provide a more geographically representative view of the health impacts of climate change. For example, almost all studies at the subnational level have so far focused on communities in high-income countries (n = 6 of 7). These biases represent the research community behind these efforts, which are almost entirely led out of Global North institutions — even when focused on health impacts in the Global South (**Figure S2**). The authorship dynamics in this subfield are not atypical in global health, which suffers from a deeply inequitable and colonial system for exchanging data, knowledge, scientific credit, and international aid^39^. Some researchers have suggested that surfacing more open access health datasets from governments in the Global South will help close this gap^5^, but this is at best a partial solution, and at worst will reinforce existing dynamics by making it easier for Global North researchers to bypass collaboration altogether. The best way to increase knowledge about the health impacts of climate change is to increase investment in research led by climate scientists and public health researchers living at the frontlines of climate injustice: as one expert recently observed, “knowledge from the global South is in the global South”^40^.

## Online Methods

### Search strategy

In this study, we focus on studies that conduct end-to-end health impact attribution, which we define as a statistical analysis that quantifies present-day or historical health impacts or risks resulting from anthropogenic (human-caused) forcings on the climate through the comparison of factual and counterfactual scenarios (where the counterfactual scenario usually excludes all anthropogenic forcings). These studies constitute some of the strongest evidence for the health impacts of climate change. However, this is a narrow definition: for example, most health outcomes that are attributable to climate change, based on the broader definition used by the Intergovernmental Panel on Climate Change^41^, have not been identified through the use of an end-to-end impact attribution study.

We used the following keyword set to search for relevant literature on the detection and attribution of human health outcomes to human-caused climate change: (“climate chang*” OR “climatic change” OR “changing climate” OR “global warming” OR “drought” OR “flood*” OR “storm*” OR “cyclone” OR “extreme weather” OR “monsoon” OR “sea level rise” OR “sea-level rise” OR “heat stress” OR “global heating”) AND (health OR mortality OR morbidity OR “infectious disease” OR “non-communicable disease” OR suicide OR stunting OR miscarriage OR diarrhea OR diarrhoea OR injuries OR cancer OR diabetes OR cardiovascular disease OR stroke OR malnutrition OR malnourish OR anxiety OR depression) AND (attribut* OR counterfactual OR “excess mortality” OR “excess cases” OR DAMIP).

We screened PubMed for studies containing these keywords anywhere in the title, abstract, or full text (search conducted July 21, 2023), and to ensure completeness, ran a second search of Web of Science for additional studies containing these keywords in the title or abstract (search conducted September 11, 2023). In total, we screened 3,677 study abstracts for broad relevance to the health impacts of climate change, and evaluated the full text of the 552 studies that passed the first round of screening. This led to the identification of only five eligible studies (**Figure S1**). However, given the lack of standardized language across studies, which we found limited the completeness of the systematic search, we also searched Google Scholar using *ad hoc* combinations of keywords related to climate change, health, and attribution. We also searched five preprint servers (arXiv, medRxiv, bioRxiv, ResearchSquare, and SSRN) using the same *ad hoc* approach. In total, this approach led to the identification of thirteen eligible studies in our original systematic search^5^, including one that was originally excluded in a prior version of the search^15^, based on a narrower definition of health impacts.

Since the conclusion of our original systematic literature search, we have continued to monitor Google Scholar and preprint servers for newly published or preprinted studies, and have periodically added new studies to an open-access database of health impact attribution literature called the Health Attribution Library (HAL; healthattribution.org). Here, we include all seven additional publications and preprints that were posted by December 31, 2024.

### Inclusion criteria

We identified a total of 20 peer-reviewed publications and preprints that have conducted end-to-end attribution of human health outcomes to human-caused climate change. These studies all include (1) a statistical model that relates health outcomes to climate variables, and (2) an estimate of the health impact of human-caused climate change, isolated through the use of a counterfactual scenario that captures natural climate variability without anthropogenic forcings. We excluded studies that quantified health effects of observed climate change by comparing health outcomes at different points in time (e.g.,^42,43^), rather than comparing present-day impacts to a present-day counterfactual that omitted anthropogenic influence on the climate. (However, we included one study that compared present-day temperature anomalies, baselined on 1880-1910, to a “pre-industrial” climate defined as an anomaly of +0°C.^44^) We also excluded a small number of studies that used present-day counterfactual climate scenarios that did not sufficiently distinguish natural and anthropogenic sources of variability^45,46^. Finally, we excluded studies that included a health-related analysis alongside a climate change attribution component, but stopped short of end-to-end attribution of health impacts^47^.

### Value of statistical life analyses

Following Newman and Noy^21^, we conducted a secondary analysis of economic losses based on the studies in Table S1. Like Newman and Noy, we use the value of statistical life to assess the economic value of deaths attributable to human-caused climate change. Such an approach monetizes lives lost by leveraging estimates of how individuals make their own tradeoffs between income (e.g., a higher-paying job) and mortality risk (e.g., a riskier job). Resulting calculations reflect the willingness of individuals to pay to avoid the elevated risk of death, and are widely used for policy evaluation in the U.S. and internationally^48^. However, such a valuation approach does not account for any broader societal economic losses, such as costs to a public healthcare system induced by elevated morbidity or mortality.

To implement this, we follow Newman and Noy in using the U.S. Environmental Protection Agency’s value for the VSL, and we treat VSL as equivalent across countries on equity-related grounds. However, we adjust the EPA figure for inflation (US $7.4m in 2006 = US $11.5m in 2024). We also present two options for how to adjust VSL estimates derived by the EPA for the U.S. population to global populations. In option 1, we adjust for the difference between U.S. and global GDP per capita (World Bank estimates: US $81,695 versus US $13,138) using an income elasticity of the VSL of 1, reflecting recent economic consensus^48^. This approach reduces our VSL estimate to US $1.85m. In option 2 (which we present in parentheticals throughout), we use the full U.S.-based value everywhere, as Newman and Noy did.

### Estimating climate change-attributable mortality from vector-borne diseases

To supplement the three studies that generated global, cause-specific estimates of climate change-attributable mortality^11,21^, we also extrapolated mortality resulting from two vector-borne diseases, based on preliminary results from two preprints^12,19^. For dengue fever, we used Childs *et al*.’s estimate that 18% (95% CI: 11% – 27%) of present-day (1995-2014) cases are due to climate change. We multiplied this proportion by an estimated 21,803 (95% CI: 17,408 – 26,030) deaths per year due to dengue fever over the same time frame, derived from the Global Burden of Disease study database^49^. For malaria, we used Carlson *et al*.’s estimate that climate change was responsible for 0.09 (95% CI: −0.30 – 0.51) percentage points of the prevalence of malaria in children (ages 2 to 10) in sub-Saharan Africa between 2010 and 2014, compared to an estimated continent-wide prevalence of 24% in 2015^50^. In the absence of an epidemiological model relating changes in childhood malaria prevalence to age-structured incidence dynamics (and ideally, accounting for interventions^51^), we assumed changes in prevalence reflect proportional changes in mortality, and used an estimate of 631,000 total deaths (95% CI: 394,000 – 914,000) across ages and across the continent in 2015^52^.

### Display Items

**Table 1.** Major estimates of global cause-specific mortality from climate change. See Online Methods for methods used in derivation of vector-borne disease mortality estimates.

| Type | Study (time period) | Cause | Deaths (annual) |
| --- | --- | --- | --- |
| Attribution studies | Carlson *et al.*<sup>19</sup> (2010 to 2015) | Malaria | 2,366<br>(95% CI: -4,925 – 19,423) |
|  | Newman & Noy<sup>21</sup> (2000 to 2019) | Extreme weather (excl. heat waves) | 2,560 |
|  | Childs *et al.*<sup>12</sup> (1995 to 2014) | Dengue fever | 3,925<br>(95% CI: 1,915 – 7,028) |
|  | Vicedo-Cabrera *et al.*<sup>11</sup> (1991 to 2018) | Heat (*in 43 countries) | 9,702<br>(95% CI: 4,005 – 19,135) |
|  | Park *et al.*[Citation error] (2010 to 2019) | Fire-related air pollution mortality | 12,566<br>(range: 3,481 – 30,126) |
|  | All attribution studies | **Total** | **31,119 deaths per year** |
| Projection studies | McMichael *et al.*<sup>27</sup> (2000) | Floods | 2,000 |
|  |  | Heat | 12,000 |
|  |  | Malaria | 27,000 |
|  |  | Diarrheal disease | 47,000 |
|  |  | Malnutrition | 77,000 |
|  |  | **Total** | **166,000 deaths per year** |
|  | Hales *et al.*<sup>29</sup> (2030) | Dengue fever | 258<br>(range: 136 to 331) |
|  |  | Heat (> 65 years old) | 37,588<br>(range: 26,912 – 48,390) |
|  |  | Diarrheal disease (< 15 years old) | 48,114<br>(range: 21,097 – 67,702) |
|  |  | Malaria | 60,091<br>(range: 37,608 – 117,001) |
|  |  | Undernutrition (< 5 years old) | 95,176<br>(range: -119,807 – 310,156) |
|  |  | **Total** | **241,227 deaths per year** |

#### Box 1. Health impact attribution and litigation.

Courts are increasingly hearing cases that aim to hold major emitters liable for their contribution to the impacts of climate change. Health impacts are often at the heart of this litigation – and health impact attribution studies are becoming an important source of potential evidence.

Many of these suits seek financial compensation for high-emitting companies’ contributions to costs incurred by climate change impacts on human health (see e.g., *Luciano Lliuya v. RWE*, where a Peruvian Ministry of Health assessment of flood risk experienced by the plaintiff was cited; or *County of Multnomah v. Exxon Mobil Corp*., where plaintiffs sought damages for “impacts on public health and infrastructure”). In these lawsuits, claimants may need to demonstrate a causal link between firms’ greenhouse gas emissions and the impacts on plaintiffs, a legal need that may be met through reference to the findings of attribution studies^25^. For example, in May 2024, three NGOs brought a criminal complaint grounded in attribution science that argued that Total Energies’ directors and main shareholders should be held criminally liable for endangering lives on the basis of the company’s contribution to climate change. Cases like these could deliver justice for victims of climate change, and discourage continued harmful conduct if firms are held liable for the costs of their emissions or directors are held personally responsible^53^.

Other cases seek accelerated mitigation action. In the 2024 ruling of the European Court of Human Rights in *Verein Klimaseniorinnen Schweiz and others v. Switzerland*, the Court referred to attribution findings in finding evidence of a causal relationship between greenhouse gas emissions and the risk of heat wave-related deaths; this assessment was necessary for the claimants to have standing to bring the case. The Court then ruled that the Swiss government’s climate policy was insufficient to protect the claimant’s rights under the European Convention. Switzerland is now forced to reassess the ambition of their climate policy in light of this ruling — or else, be found to continue to be in breach of the European Convention on Human Rights.

## Supplemental Figures and Tables

**Figure S1.**
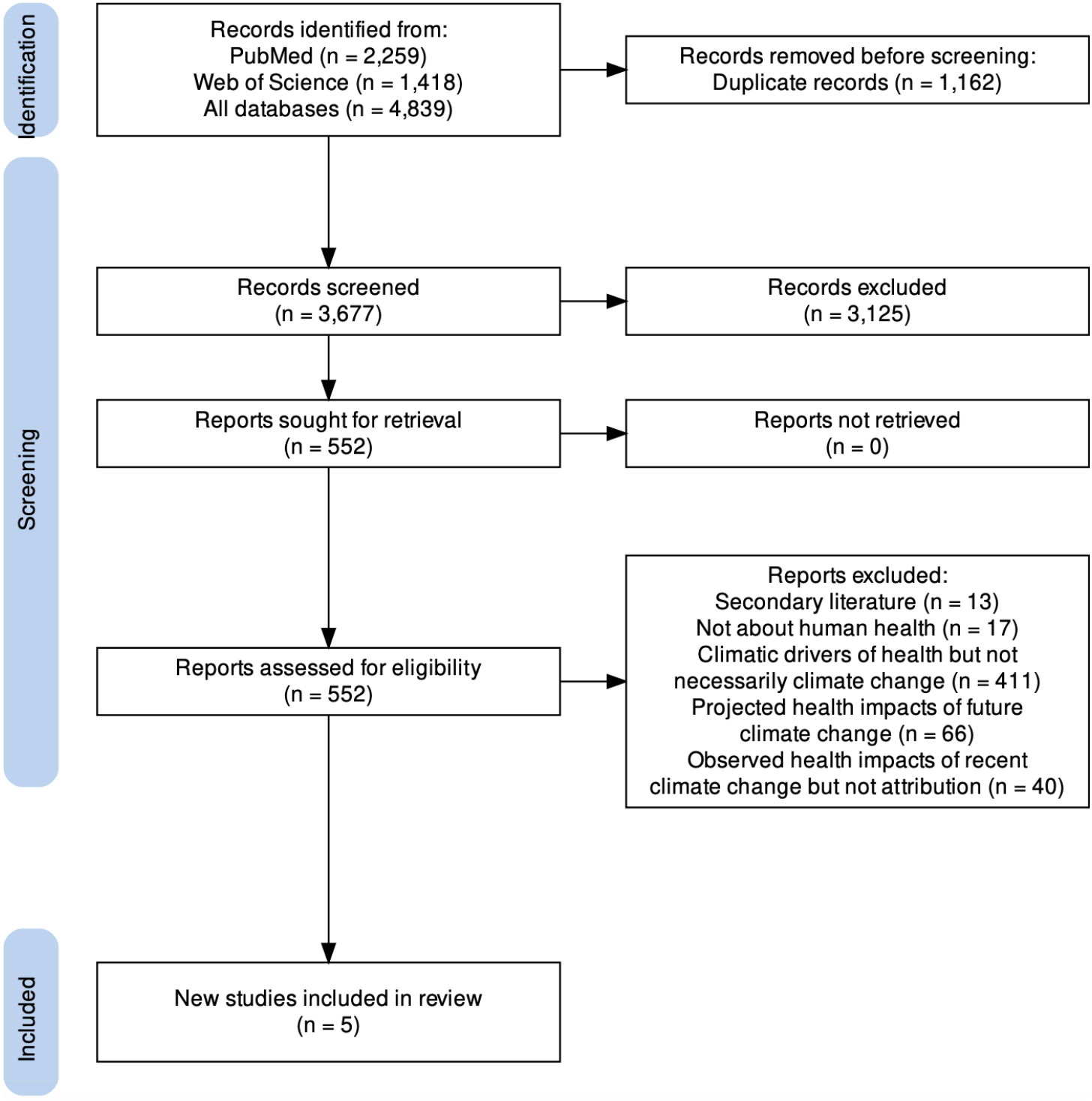
Results of the systematic literature review. The PRISMA template was generated by the PRISMA2020 R Shiny app^54^.

**Figure S2.**
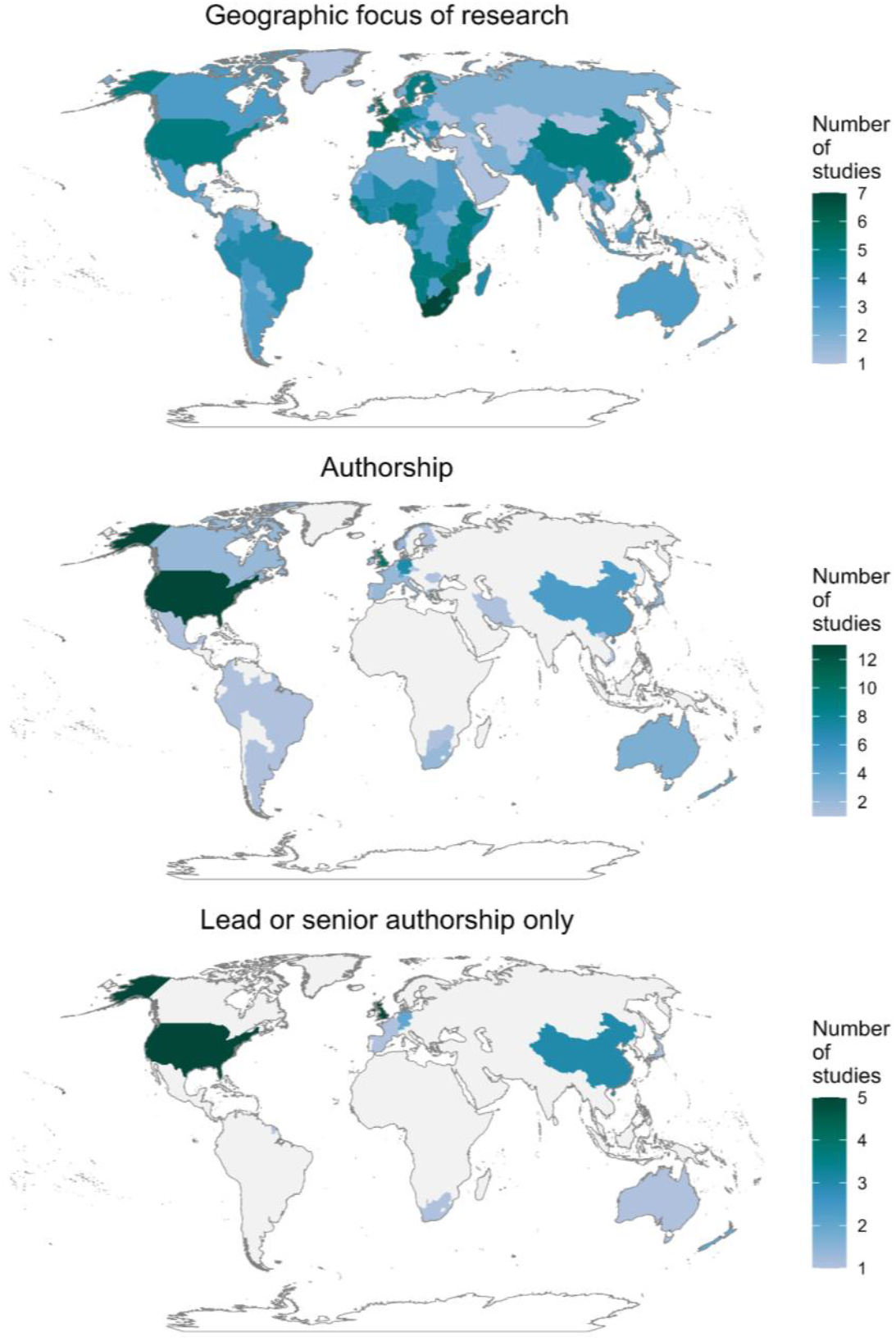
Geography of study scope and author affiliations. Gray values indicate no studies were applicable. (Middle and bottom panel indicate the number of studies with at least one author, or lead author, with an affiliation listed in that country.)

**Table S1.** Estimates of deaths attributed to anthropogenic climate change. Estimates that are italicized are preliminary findings from preprints, and so may be subject to change. Only estimates of attributable mortality are shown, and not estimates of deaths averted^7,16,21^.

| Cause, location, and period | Estimate |
| --- | --- |
| Heat-related deaths from the hottest day of the summer 2006 heat wave in London, UK (July 26, 2006)<sup>8</sup> | 10 deaths |
| Deaths due to Hurricane Harvey in Texas, USA (2017) (extrapolated)<sup>20</sup> | 51 – 66 deaths |
| Heat-related deaths due to the summer 2003 heat wave in Greater London, UK (June 1 to August 31, 2003)<sup>6</sup> | 64 deaths<br>(95% CI: 61 – 67) |
| *Warm season heat-related deaths during the summer 2018 heat wave in the Canton of Zürich, Switzerland (July 29 to August 10, 2018)<sup>9</sup>* | *86 deaths*<br>*(95% CI: 55 – 113)* |
| Heat-related deaths during the summer 2022 heat wave in Switzerland (June 1 to August 31, 2022)<sup>10</sup> | 370 deaths<br>(95% CI: 133 – 644) |
| Heat-related deaths due to the summer 2003 heat wave in Central Paris, France (June 1 to August 31, 2003)<sup>6</sup> | 506 deaths<br>(95% CI: 455 – 557) |
| Neonatal deaths related to preterm births in China (2010 to 2020)<sup>17</sup> | 682 deaths<br>(95% CI: 99 – 902) |
| Warm season heat-related neonatal death due to pre-term births in China (2010 to 2020)<sup>17</sup> | 1,276 deaths<br>(95% CI: 924 – 1,881) |
| *Warm season heat-related deaths in the Canton of Zürich, Switzerland (1969 to 2018)<sup>9</sup>* | *1,683 deaths*<br>*(95% CI: 270 – 3,279)* |
| Year-round heat-related childhood deaths in Africa (1995 to 2004)<sup>55</sup> | ~3,000 – ~5,000 deaths |
| Global deaths due to PM2.5 air pollution from fires (1960 to 1969)<sup>14</sup> | 6,690 deaths<br>(range: -3,980 – 17,730) |
| Year-round heat-related childhood deaths in Africa (2011 to 2020)<sup>55</sup> | ~7,000 – ~11,000 deaths |
| Global deaths due to 154 extreme weather events (heat and cold waves, floods and droughts, storms, and wildfires) that became more likely due to climate change (2000 to 2019)<sup>21</sup> | 75,139 deaths |
| Global deaths due to PM2.5 air pollution from fires (2010 to 2019)<sup>14</sup> | 125,660 deaths<br>(range: 34,810 – 301,260) |
| Global heat-related neonatal deaths (2001 to 2019)<sup>16</sup> | 175,133 deaths<br>(95% CI: 7,806 – 353,516) |
| Global warm season heat-related deaths across 732 populations from 43 countries (seasonal average summed over 1991 to 2018)<sup>11</sup> | 271,656 deaths<br>(95% CI: 112,140 – 535,780) |

## Author Contributions

CJC and DM designed the study. CJC, RJG, TEL, CAL, MLS, MAN, SJR, and DSS generated data. CJC, RJG, and TAC contributed to data analysis and data visualization. CJC, RFSS, and TAC drafted the manuscript, and all authors edited and approved the final manuscript.

## Data Availability

Statement All data and code are publicly available at github.com/carlsonlab/AttributableLosses.

## Acknowledgments

We thank Felipe J. Colon-Gonzalez and Christopher Callahan for thoughtful conversations about health impact attribution. The authors declare no conflict of interest.

## References

1. Forster, P. M. et al. Indicators of Global Climate Change 2023: annual update of key indicators of the state of the climate system and human influence. Earth Syst. Sci. Data 16, 2625–2658 (2024).

2. Carlson, C. J. et al. Designing and describing climate change impact attribution studies: A guide to common approaches. earthRxiv (2024) doi :10.31223/X5CD7M.

3. Ebi, K. L., Ogden, N. H., Semenza, J. C. & Woodward, A. Detecting and Attributing Health Burdens to Climate Change. Environ. Health Perspect. 125, 085004 (2017).

4. Ebi, K. L. et al. Using Detection And Attribution To Quantify How Climate Change Is Affecting Health. Health Aff. 39, 2168–2174 (2020).

5. Carlson, C. J. et al. Detection and Attribution of Climate Change Impacts on Human Health: A Data Science Framework. Wellcome Open Research 9, 245 (2024).

6. Mitchell, D. et al. Attributing human mortality during extreme heat waves to anthropogenic climate change. Environ. Res. Lett. 11, 074006 (2016).

7. Hajat, S., Gampe, D. & Petrou, G. Contribution of Cold Versus Climate Change to Mortality in London, UK, 1976–2019. Am. J. Public Health 114, 398–402 (2024).

8. Perkins-Kirkpatrick, S. E. et al. On the attribution of the impacts of extreme weather events to anthropogenic climate change. Environ. Res. Lett. 17, 024009 (2022).

9. Stuart-Smith, R. et al. Quantifying heat-related mortality attributable to human-induced climate change. Research Square (2023) doi :10.21203/rs.3.rs-2702337/v2.

10. Vicedo-Cabrera, A. M. et al. The footprint of human-induced climate change on heat-related deaths in the summer of 2022 in Switzerland. Environ. Res. Lett. 18, 074037 (2023).

11. Vicedo-Cabrera, A. M. et al. The burden of heat-related mortality attributable to recent human-induced climate change. Nat. Clim. Chang. 11, 492–500 (2021).

12. Childs, M. L., Lyberger, K., Harris, M., Burke, M. & Mordecai, E. A. Climate warming is expanding dengue burden in the Americas and Asia. medRxiv (2024) doi :10.1101/2024.01.08.24301015.

13. Erazo, D. et al. Contribution of climate change to the spatial expansion of West Nile virus in Europe. Nat. Commun. 15, 1196 (2024).

14. Park, C. Y. et al. Attributing human mortality from fire PM2.5 to climate change. Nat. Clim. Chang. 14, 1193–1200 (2024).

15. Mester, B. et al. Human displacements from Tropical Cyclone Idai attributable to climate change. Nat. Hazards Earth Syst. Sci. 23, 3467–3485 (2023).

16. Dimitrova, A. et al. Temperature-related neonatal deaths attributable to climate change in 29 low- and middle-income countries. Nat. Commun. 15, 5504 (2024).

17. Zhang, Y. et al. The burden of heatwave-related preterm births and associated human capital losses in China. Nat. Commun. 13, 7565 (2022).

18. Zhu, Z. et al. Estimating the burden of temperature-related low birthweight attributable to anthropogenic climate change in low-income and middle-income countries: a retrospective, multicentre, epidemiological study. Lancet Planet. Health 8, e997–e1009 (2024).

19. Carlson, C. J., Carleton, T. A., Odoulami, R. C. & Trisos, C. H. The historical fingerprint and future impact of climate change on childhood malaria in Africa. bioRxiv (2023) doi :10.1101/2023.07.16.23292713.

20. Frame, D. J., Wehner, M. F., Noy, I. & Rosier, S. M. The economic costs of Hurricane Harvey attributable to climate change. Clim. Change 160, 271–281 (2020).

21. Newman, R. & Noy, I. The global costs of extreme weather that are attributable to climate change. Nat. Commun. 14, 6103 (2023).

22. Kniesner, T. J. & Viscusi, W. K. The value of a statistical life. Oxford Research Encyclopedia of Economics and Finance (2019) doi :10.1093/acrefore/9780190625979.013.138.

23. Noy, I. et al. Event attribution is ready to inform loss and damage negotiations. Nat. Clim. Chang. 1–3 (2023).

24. US Environmental Protection Agency. Report on the social cost of greenhouse gases: Estimates incorporating recent scientific advances. https://www.epa.gov/system/files/documents/2023-12/epa_scghg_2023_report_final.pdf (2022).

25. Stuart-Smith, R. F. et al. Filling the evidentiary gap in climate litigation. Nat. Clim. Chang. 11, 651–655 (2021).

26. Burger, M., Wentz, J. & Horton, R. The law and science of climate change attribution. Colum. J. Envtl. L. 45, 57 (2020).

27. McMichael, A. J. et al. Climate Change. in Chapter 20. Comparative Quantification of Health Risks (World Health Organization (WHO Press), 2004).

28. Carlson, C. J. After millions of preventable deaths, climate change must be treated like a health emergency. Nat. Med. (2024) doi :10.1038/s41591-023-02765-y.

29. World Health Organization. Quantitative risk assessment of the effects of climate change on selected causes of death, 2030s and 2050s. (2014). World Health Organization: Geneva, Switzerland.

30. Dasgupta, S. & Robinson, E. J. Z. Attributing changes in food insecurity to a changing climate. Sci. Rep. 12, 4709 (2022).

31. Levy, K., Woster, A. P., Goldstein, R. S. & Carlton, E. J. Untangling the impacts of climate change on waterborne diseases: A systematic review of relationships between diarrheal diseases and temperature, rainfall, flooding, and drought. Environ. Sci. Technol. 50, 4905–4922 (2016).

32. Wang, P., Asare, E., Pitzer, V. E., Dubrow, R. & Chen, K. Associations between long-term drought and diarrhea among children under five in low- and middle-income countries. Nat. Commun. 13, 3661 (2022).

33. Wang, P., Asare, E. O., Pitzer, V. E., Dubrow, R. & Chen, K. Floods and diarrhea risk in young children in low- and middle-income countries. JAMA Pediatr. 177, 1206–1214 (2023).

34. Perkins-Kirkpatrick, S. E. et al. Attributing heatwave-related mortality to climate change: a case study of the 2009 Victorian heatwave in Australia. Environ. Res.: Climate 4, 015004 (2025).

35. Callahan, C. et al. Quantifying the contributions of climate change and adaptation to mortality from unprecedented extreme heat events. EarthArXiv (2025) doi :10.31223/x51x5x.

36. Ebi, K. L. et al. Estimating the total effect of anthropogenic climate change on human health and wellbeing. Nat. Med. (2024) doi :10.1038/s41591-024-03051-1.

37. Rother, H.-A. Controlling and preventing climate-sensitive noncommunicable diseases in urban sub-Saharan Africa. Sci. Total Environ. 722, 137772 (2020).

38. Carleton, T. A. Crop-damaging temperatures increase suicide rates in India. Proc. Natl. Acad. Sci. U. S. A. 114, 8746–8751 (2017).

39. Abimbola, S. The foreign gaze: authorship in academic global health. BMJ Glob Health 4, e002068 (2019).

40. Abimbola, S. Knowledge from the global South is in the global South. J. Med. Ethics 49, 337–338 (2023).

41. Hegerl, G. C. et al. Good practice guidance paper on detection and attribution related to anthropogenic climate change. (2010). World Meteorological Organization: Geneva, Switzerland.

42. Oudin Åström, D., Forsberg, B., Ebi, K. L. & Rocklöv, J. Attributing mortality from extreme temperatures to climate change in Stockholm, Sweden. Nat. Clim. Chang. 3, 1050–1054 (2013).

43. Silva, R. A. et al. Global premature mortality due to anthropogenic outdoor air pollution and the contribution of past climate change. Environ. Res. Lett. 8, 034005 (2013).

44. Beck, T. et al. Increasing Likelihood of Heat-Related Mortality Events with Global Warming: A Continental Epidemiological Extreme Event Attribution Study. Research Square (2024) doi:10.21203/rs.3.rs-5549732/v1.

45. Alonso, D., Bouma, M. J. & Pascual, M. Epidemic malaria and warmer temperatures in recent decades in an East African highland. Proc. Biol. Sci. 278, 1661–1669 (2011).

46. Head, J. R. et al. Effects of precipitation, heat, and drought on incidence and expansion of coccidioidomycosis in western USA: a longitudinal surveillance study. Lancet Planet Health 6, e793–e803 (2022).

47. Harris, M. J. et al. Extreme precipitation, exacerbated by anthropogenic climate change, drove Peru’s record-breaking 2023 dengue outbreak. medRxiv (2024) doi :10.1101/2024.10.23.24309838.

48. Viscusi, W. K. The Role of Publication Selection Bias in Estimates of the Value of a Statistical Life. American Journal of Health Economics 1, 27–52 (2015).

49. GBD 2021 Causes of Death Collaborators. Global burden of 288 causes of death and life expectancy decomposition in 204 countries and territories and 811 subnational locations, 1990-2021: a systematic analysis for the Global Burden of Disease Study 2021. Lancet 403, 2100–2132 (2024).

50. Snow, R. W. et al. The prevalence of Plasmodium falciparum in sub-Saharan Africa since 1900. Nature 550, 515–518 (2017).

51. Bhatt, S. et al. The effect of malaria control on Plasmodium falciparum in Africa between 2000 and 2015. Nature 526, 207–211 (2015).

52. Gething, P. W. et al. Mapping Plasmodium falciparum mortality in Africa between 1990 and 2015. N. Engl. J. Med. 375, 2435–2445 (2016).

53. Wetzer, T., Stuart-Smith, R. & Dibley, A. Climate risk assessments must engage with the law. Science 383, 152–154 (2024).

54. Haddaway, N. R., Page, M. J., Pritchard, C. C. & McGuinness, L. A. PRISMA2020: An R package and Shiny app for producing PRISMA 2020-compliant flow diagrams, with interactivity for optimised digital transparency and Open Synthesis. Campbell Syst Rev 18, e1230 (2022).

55. Chapman, S. et al. Past and projected climate change impacts on heat-related child mortality in Africa. Environ. Res. Lett. 17, 074028 (2022).

